# SARS-CoV-2 infections among pregnant women, 2020, Finland – cross-testing of neutralization assays

**DOI:** 10.1101/2023.06.07.23290927

**Authors:** J Virtanen, EM Korhonen, S Salonen, O Vapalahti, T Sironen, AJ Jääskeläinen

**Author notes:** Corresponding author: Jenni Virtanen.

## Abstract

We studied the development of SARS-CoV-2 pandemic in Finland in 2020 and evaluated the performance of two surrogate immunoassays for detection of neutralizing antibodies (NAbs). The dataset consisted of 12000 retrospectively collected samples from pregnant women in their 1^st^ trimester throughout 2020. All the samples were initially screened for IgG with SARS-CoV-2 spike antibody assay (EIM-S1, Euroimmun, Lübeck, Germany) followed by confirmation with nucleocapsid antibody assay (Architect SARS-CoV-2, Abbott, Illinois, USA). Samples that were reactive (positive or borderline) with both assays were subjected to testing with commercial surrogate immunoassays of NeutraLISA (EIM) and cPass^TM^ (GenScript Biotech Corporation, Rijswijk, Netherlands) by using pseudoneutralization assay (PNAbA) as a golden standard. No seropositive cases were detected between January and March. Between April and December, IgG (EIM-S1 and Abbott positive) and NAb (PNAbA positive) seroprevalences were between 0.4-1.4%. NeutraLISA showed 90% and cPass 55% concordant results with PNAbA among PNAbA negative samples and 49% and 92% among PNAbA positive samples giving NeutraLISA better specificity but lower sensitivity than cPass. To conclude, seroprevalence in pregnant women reflected that of the general population but the variability of the performance of serological protocols needs to be taken into account in inter-study comparison.

## Introduction

The first case of severe acute respiratory syndrome coronavirus 2 (SARS-CoV-2) in Finland was reported in a traveler from Wuhan late January 2020 [1]. The first endemic peak emerged late March and early April after which the case numbers slowly declined before a new wave late summer 2020 [2]. Preventive measures included national gathering restrictions, online schooling, and mask recommendations/requirements as well as isolation of the most affected Helsinki and Uusimaa region from late March until mid-April 2020. Based on antibody screening by the Finnish institute for Health and Welfare (THL), monthly seroprevalence in the community of Helsinki and Uusimaa hospital district (HUS) between April and December 2020 ranged from 0.3% to 5.5% reaching its peak in November, while the prevalence in the rest of the country remained lower [3]. The vaccination campaign begun with healthcare workers at the end of December 2020 and continued during 2021 with other population groups.

Serological assays detecting IgG are commonly used in epidemiological surveys and population level monitoring [4]. Only part of the antibodies produced can neutralize the virus itself, and further protect the individual, however, other antibodies may help to protect the individual *in vivo* [5]. Monitoring of neutralizing antibodies (NAbs) instead of IgG could be used, for example, to follow the level of protective immunity of immunosuppressed patients or candidates for convalescent plasma therapy or to assess the success of vaccination. Plaque-reduction test and neutralization test or its pseudovirus-based applications are commonly used as a golden standard for NAb detection but are mainly utilized for research purposes because of their complexity and requirements for high-containment facilities [6–8]. Due to this, several commercial surrogate assays for detecting NAbs have been designed. However, they have displayed discrepant performances in different laboratories [4, 6, 8, 9]. Performance of different immunoassays, including the ones detecting NAbs originating from natural infection or vaccination, should be validated carefully in order to be used in diagnostic settings for, e.g., re-vaccination purposes. As post-vaccination antibodies can be high and affect the results in assays detecting NAbs, e.g., by increasing sensitivity [7], we wanted to test these assays after natural infection without vaccination history. Surrogate assays that show optimal performance with convalescent sera should then be further validated with individuals with vaccination or hybrid immunity.

Here, we screened antibodies against SARS-CoV-2 in 12000 pregnant women sampled in routine maternity screening between January and December 2020 to follow the development of the pandemic in Finland and to evaluate the infection risk to expectant mothers as well as the performance of commercial surrogate assays designed for detection of NAbs. Initial screening was carried out with sensitive commercial immunoassay detecting SARS-CoV-2 spike antibodies (EIM), followed by a more specific immunoassay detecting antibodies against SARS-CoV-2 nucleocapsid (Abbott). The reactive samples were tested for Nabs using a pseudoneutralization assay (PNAbA; [10]). Finally, two different commercial surrogate immunoassays of NeutraLISA (EIM) and cPass^TM^ (GenScript), Rijswijk, Netherlands] aimed for detection of SARS-CoV-2 NAbs were validated against PNAbA.

## Methods

### Sampling and dataset

Altogether, 12000 serum samples were retrospectively collected in 2020 (1000 samples per month). These serum samples were originally sent for routine screening of hepatitis B S-antigen, antibodies and antigen for HIV, and syphilis antibodies during the first trimester of pregnancy (Helsinki University Hospital, Diagnostic center, Helsinki, Finland). Samples were randomly collected and re-coded. Only age, hospital district, and SARS-CoV-2 PCR info were available. Ethics committee and institutional review board of Helsinki University Hospital gave ethical approval for this work (HUS/32/2018, HUS/157/2020, HUS/151/2022, HUS/244/2021, HUS/32/2018). All the samples were treated as anonymous according to research permits.

Samples originated from three hospital districts in Southern Finland: Hospital District of Helsinki and Uusimaa (HUS), South Karelia Social and Health Care District (Eksote), and Social and Health Services in Kymenlaakso (Kymsote) (Fig. 1). Together, these areas inhabit 36% of Finnish population (Table 1 and S1) and mean population density is 106 inhabitants/km^2^ [11]. During 2020, first COVID-19 cases were reported Jan 29^th^ and Feb 26^th^ (travelers) and by the end of the year, 1.14% of the population in study areas had had a laboratory confirmed disease, the highest number being in the most densely populated HUS region [2].

**Fig. 1.**
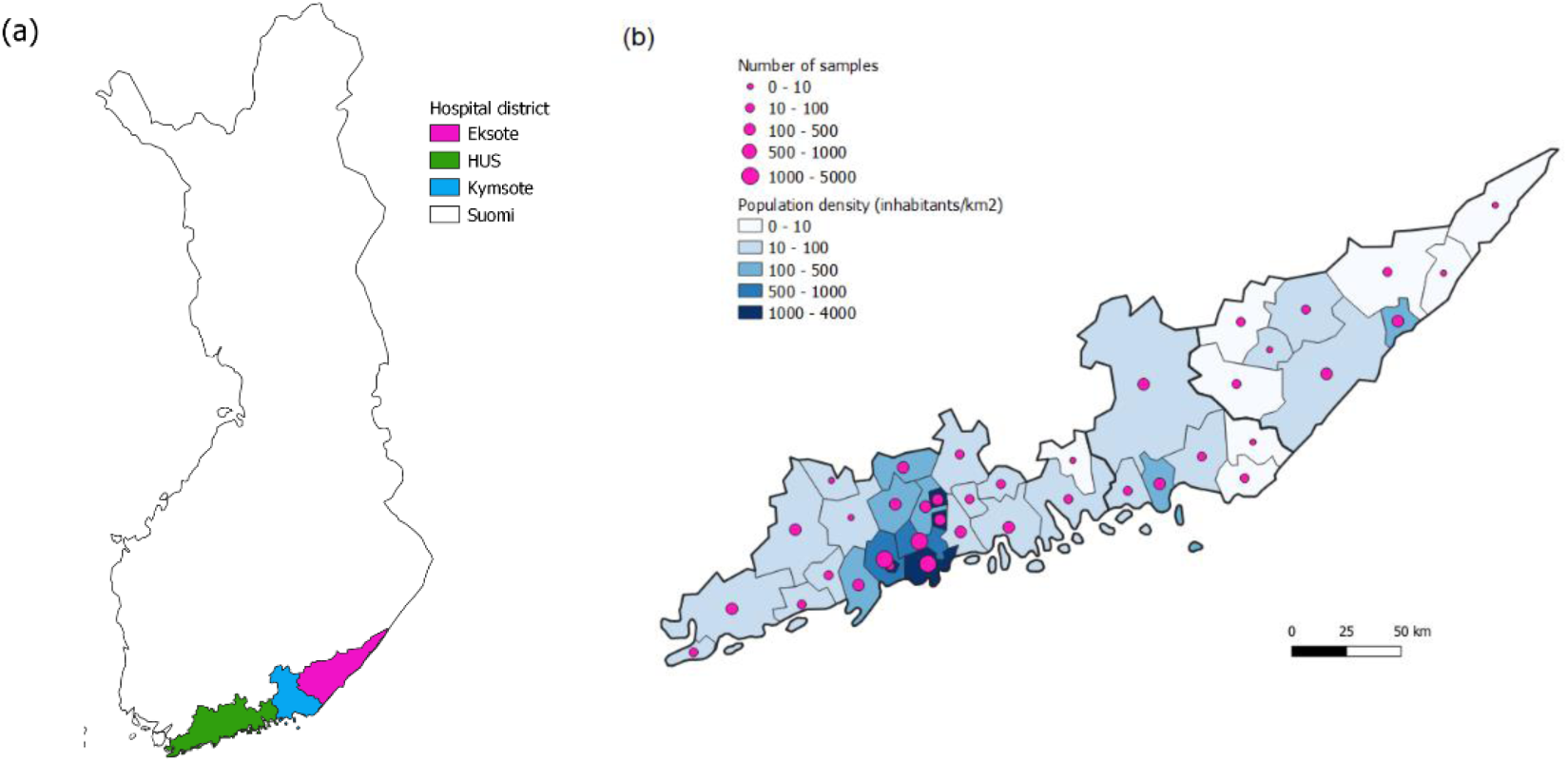
Location of the hospital districts (a) and sample amounts and population densities in the municipalities in the region (b).

**Table 1.** Geographical information about hospital districts and population

| Hospital district | Samples (n) | Inhabitants (n) | Inhabitants (% from Finnish population) | Population density (inhabitants/km2) |
| --- | --- | --- | --- | --- |
| Eksote | 493 | 127757 | 2 | 24 |
| Kymsote | 663 | 164456 | 3 | 36 |
| HUS | 10844 | 1685983 | 31 | 193 |
| Total | 12000 | 1978196 | 36 | 106 |

### Screening of SARS-CoV-2 spike and N IgG antibodies

All 12000 serum samples were screened for IgG antibodies with EUROLabworkstation (EIM) and SARS-CoV-2 IgG kit (S1-based antigen, EIM; later EIM-S1-assay) according to manufacturer’s instructions. Samples that were reactive with the EIM-S1-assay (result positive or borderline, ≥0.8 index) were subjected to a second screening using Architect Analyzer (Abbott, Illinois, USA) and SARS-COV-2 IgG (nucleoprotein-based antigen, Abbott; later Abbott-N-assay) according to manufacturerś instructions (Table S2). All the samples that were ≥0.8 index (manufacturer reported limit for borderline result) with EIM-S1-assay and ≥0.3 index with Abbott-N-assay (borderline limit determined based on earlier experience (data not shown) and this study (Fig. S1)) were considered reactive and were further selected for NAb screening. Also, individuals who had been previously positive in SARS-CoV-2 nucleic acid test were selected for NAb testing (Table S2).

### PNAbA and commercial surrogate ELISAs for detection of SARS-CoV-2 neutralizing antibodies

NAbs were analyzed with PNAbA described earlier [10, 12, 13] (University of Helsinki, Helsinki, Finland) and two commercial surrogate immunoassays: SARS-CoV-2 NeutraLISA (EIM) and cPass^TM^ SARS-CoV-2 Neutralization Antibody Detection Kit (GenScript). Both commercial surrogate assays were carried out and interpreted according to manufactureŕs instructions. Both surrogate assays detect neutralizing antibodies, which inhibit the binding of S1/RBD to ACE2-reseptors [14]. According to the manufacturers, NeutraLISA and cPass have sensitivity of 95.9% and specificity 99.7%, and 93.8% and 99.4%, respectively.

### Statistical analysis

In order to avoid false positive results, a two-tier approach was used to determine SARS-CoV-2 IgG positivity for seroprevalence calculations: those patients that were positive with both EIM-S1 (>1.1 index) and Abbott-N (>1.4 index) according to manufacturer’s guidelines, were considered positive. Borderline results were considered negative. For NAb seroprevalence, the samples were considered positive if they had a detectable titre in PNAbA (20 or above). As it was not possible to test all the 12000 samples with PNAbA, the samples which were not reactive with both IgG ELISAs (EIM-S1 and Abbott-N-assays) were considered PNAbA negative for calculation purposes. The titres below 20 were set to 10 for calculations. The 95% confidence intervals were calculated with Wilson score method (binominal proportion). ROC curve analysis was performed to compare the methods to PNAbA with titre cut-offs 20 (also week neutralization included) or 80 (only strong neutralization included). Spearman’s rho was used to study correlation. Cohen’s Kappa analysis was used to compare the results between assays. All the statistics were carried out in IBM SPSS Statistics version 28 or RStudio utilizing dplyr, ggplot2, and reshape2 packages [15–19]. Maps were drawn with QGIS 3.26.2 utilizing datasets by World Food Programme and Statistics Finland [20, 21].

## Results

### IgG and NAb seroprevalence

In total, there was one recoded sample available per individual, 10844 of which originated from HUS, 493 from Eksote, and 663 from Kymsote. The overall mean age was 31.6 years (range 14-61, SD 5.2). Mean ages in hospital districts were 31.7 (SD 5.1) in HUS, 30.7 (SD 5.1) in Eksote, and 30.6 (SD 5.4) in Kymsote region (Table S3).

Out of the 12000 individuals, 435 were reactive (either borderline or positive) in EIM-S1-assay. Only 116 of them also had reactive results in Abbot-N-assay, and all these 116 were further tested with PNAbA for NAbs. All previously PCR-positive individuals (N=35; PCR positivity varied from 0-9 months prior serum sampling) were reactive in EIM-S1-assay, but the results with Abbott-N-assay varied: 31/35 were ≥0.3 index and 4/35 were <0.3 index. These four PCR-positive but Abbott-N-assay negative samples were also tested for NAbs. In total, 91 out of these 120 samples showed NAbs using PNAbA (titre of 20 or more), including 3 out of 4 Abbott-N negative but EIM-S1-positive individuals with PCR-confirmed previous infection.

When borderline results are considered negative, a total of 0.57% out of all the 12000 individuals (67/12000; 95% CI 0.45-0.72; EIM-S1-assay≥1.1 and Abbot-N-assay≥1.4) were IgG positive and 0.76% (91/12000; 95% CI 0.62-0.93) were NAb positive. All the samples taken between January and March were IgG and NAb negative. After April, the seroprevalence (IgG or NAb) varied between 0.4-1.4% (Figure 2 and S2, Table 2). Out of the NAb positive patients, only 28% had had a PCR confirmed disease at some point before sampling with the time from the disease varying between less than one and nine months. The rest had either been negative, been tested in private health care (data not available), or not been tested at all. Ten PCR confirmed cases were PNAbA negative and the samples were taken one to six months after the infection. Out of these, four had no detectable NAbs with cPass^TM^ or NeutraLISA either, two were positive with both of them and four only with cPass^TM^. Differences were detected between hospital districts, as only one NAb positive (0.2 %) and no IgG positive patients were detected in the least densely populated Eksote region whereas 0.61 % were IgG positive and 0.80 % NAb positive in the most populated HUS region.

**Fig. 2.**
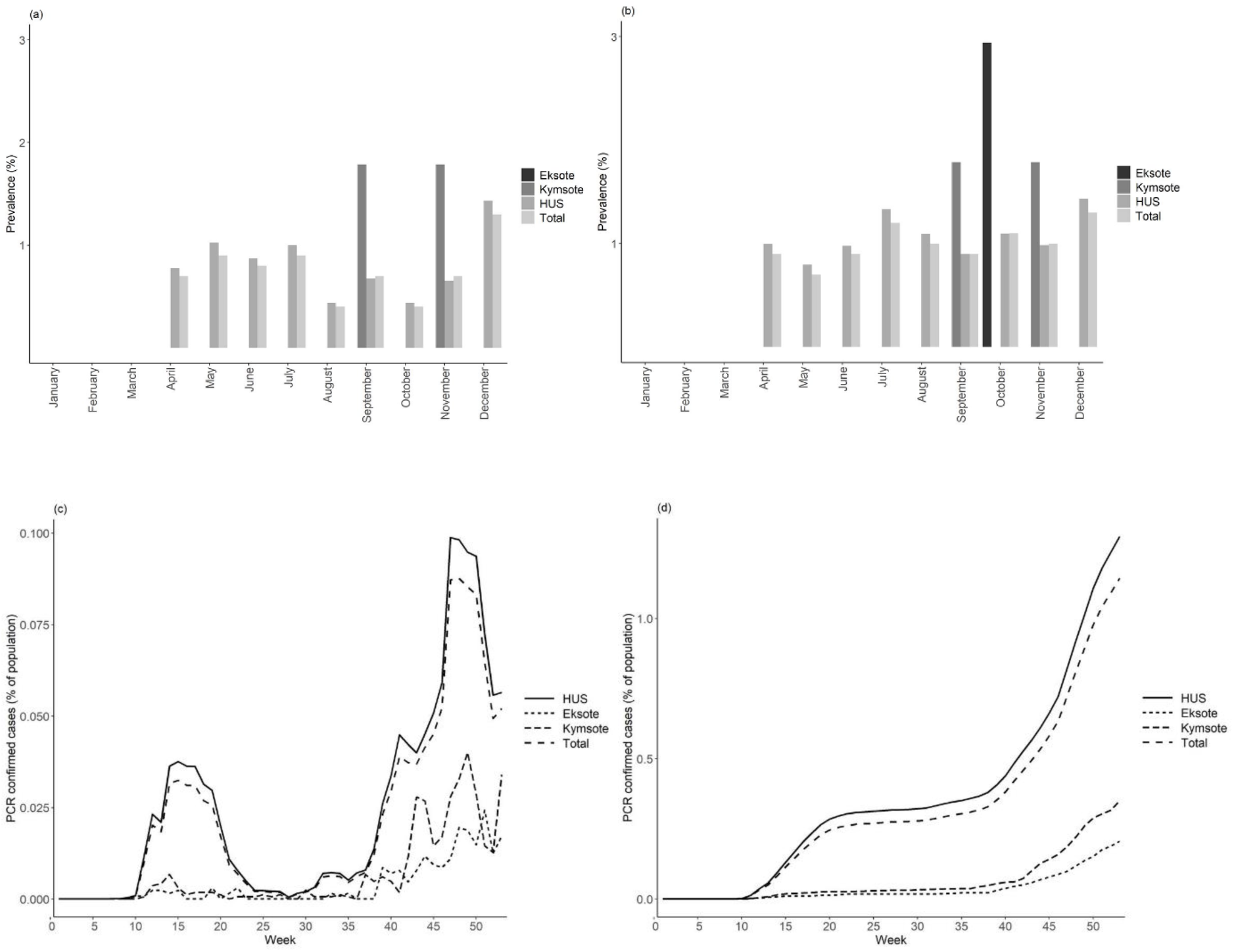
SARS-CoV-2 IgG (EIM-S1>1.1 and Abbott-N>1.4) (a) and NAb (titre 20 or over) (b) seroprevalence of this data, and reported PCR prevalence according to THL [2] (c-d) in three hospital districts in Southern Finland in 2020. Cases that were positive with both IgG tests were counted as positives for IgG and those that were positive in PNAbA as positive for NAbs. Official PCR confirmed cases in study regions (as % of whole population) are included as a comparison as weekly (c) and cumulative numbers (d).

**Table 2.** IgG and NAb prevalences by hospital district and month. Numbers are reported in parenthesis and 95% confidence intervals (Wilson score method) in brackets

|  | Positive in<br>EIM-S1 | Borderline in<br>EIM-S1 | Positive in<br>Abbott-N | IgG positive* | NAb positive | Total N |
| --- | --- | --- | --- | --- | --- | --- |
| Eksote | 1.01 (5) | 0.81 (4) | 0 (0) | 0 (0) [0.00–0.77] | 0.2 (1) [0.04–1.14] | 493 |
| Kymsote | 1.66 (11) | 1.36 (9) | 0.3 (2) | 0.3 (2) [0.08–1.09] | 0.3 (2) [0.08–1.09] | 663 |
| HUS | 2.07 (225) | 1.69 (183) | 0.65 (70) | 0.61 (66) [0.48–0.77] | 0.8 (88) [0.65–0.99] | 10844 |
| Jan | 0.6 (6) | 1.9 (19) | 0 (0) | 0 (0) [2.16E-17–0.38] | 0 (0) [2.16E-17–0.38] | 1000 |
| Feb | 0.8 (8) | 2.1 (21) | 0 (0) | 0 (0) [2.16E-17–0.38] | 0 (0) [2.16E-17–0.38] | 1000 |
| Mar | 0.8 (8) | 1.1 (11) | 0 (0) | 0 (0) [2.16E-17–0.38] | 0 (0) [2.16E-17–0.38] | 1000 |
| Apr | 1.6 (16) | 1.3 (13) | 0.9 (9) | 0.7 (7) [0.39–1.44] | 0.9 (9) [0.47–1.70] | 1000 |
| May | 1.9 (19) | 1.6 (16) | 0.9 (9) | 0.9 (9) [0.47–1.70] | 0.7 (7) [0.39–1.44] | 1000 |
| Jun | 2.4 (24) | 1.8 (18) | 0.8 (8) | 0.8 (8) [0.41–1.57] | 0.9 (9) [0.47–1.70] | 1000 |
| Jul | 2.3 (23) | 1.2 (12) | 0.9 (9) | 0.9 (9) [0.47–1.70] | 1.2 (12) [0.69–2.09] | 1000 |
| Aug | 2.4 (24) | 2.1 (21) | 0.5 (5) | 0.4 (4) [0.16–1.20] | 1.0 (10) [0.54–1.83] | 1000 |
| Sep | 2.6 (26) | 1.7 (17) | 0.7 (7) | 0.7 (7) [0.34–1.44] | 0.9 (9) [0.47–1.70] | 1000 |
| Oct | 3 (30) | 2.0 (20) | 0.4 (4) | 0.4 (4) [0.16–1.02] | 1.1 (11) [0.62–1.96] | 1000 |
| Nov | 2.8 (28) | 2.8 (13) | 0.7 (7) | 0.7 (7) [0.34–1.44] | 1.0 (10) [0.54–1.83] | 1000 |
| Dec | 2.9 (29) | 1.5 (17) | 1.4 (14) | 1.3 (13) [0.76–2.21] | 1.4 (14) [0.84–2.34] | 1000 |

\*Positive in both EIM ( $\geq 1.1$ = pos) and Abbott ( $\geq 1.4$ = pos); EIM $\geq 0.8$ -1.0 Borderline.

### Comparison of surrogate immunoassays and PNAbA

Two different commercial surrogate assays of NeutraLISA (EIM) and cPass^TM^ (GenScript) were evaluated and results compared to PNAbA (Fig. S3). In total 120 samples were tested with PNAbA. Samples with titre of 20 or more in PNAbA (N=91; Table 3) were considered as samples with true neutralizing antibodies. Of these 91 samples, NeutraLISA gave positive results in 45 samples indicating concordance of 49% and cPass^TM^ gave positive results in 84 samples with concordance of 92%. Notably, NeutraLISA gave borderline results in 22 samples positive in PNAbA. When comparing the PNAbA negative samples (N=29), NeutraLISA had 90% concordance while cPass^TM^ only 55% concordance.

**Table 3.** Comparison of PNAbA to cPass^TM^ and NeutraLISA results.

| Surrogate ELISA |  | Pseudoneutralization assay |  |  |  |
| --- | --- | --- | --- | --- | --- |
|  |  | N (120) |  | Concordant results % |  |
|  |  | Neg (29) | Pos (91) | Neg (%) | Pos (%) |
| cPass™ | Neg | 16 | 7 | 55 | 8 |
|  | Pos | 13 | 84 | 45 | 92 |
| NeutraLISA | Neg | 26 | 26 | 90 | 29 |
|  | Borderline | 1 | 20 | 3 | 22 |
|  | Pos | 2 | 45 | 7 | 49 |

Between test agreement (Cohen’s Kappa values, Table S5) ranged between slight (0.060, EIM vs Abbott) and moderate (0.511, PNAbA vs cPass^TM^). The agreement between PNAbA and NeutraLISA was 0.281. When samples with a weak titre (below 80) were considered negative for neutralizing capacity, agreement to PNAbA was fair (0.330, cPass^TM^) and substantial (0.614, NeutraLISA). All the tests showed a significant correlation with Spearman’s rho varying between 0.448 (Abbott-N and cPass^TM^) and 0.884 (NeutraLISA vs cPass^TM^, Table S4, Fig. 3 and S4). Correlation with PNAbA titre was 0.819 with cPass^TM^ and 0.757 with NeutraLISA. AUC value based on ROC curve (Figure S5, considering PNAbA a golden standard) was highest with EIM-S1 (0.882) and lowest with Abbott-N (0.758) when titre cut off 20 was used. When cut off 80 was used, cPass^TM^ had the highest AUC (0.920) and Abbott-N the lowest (0.732). However, using of manufacturer reported guidelines lead to big false positive rates in almost all the tests. Only with NeutraLISA, the sensitivity was 71% and false positivity rate only 11% when cut off 80 was used.

**Fig. 3.**
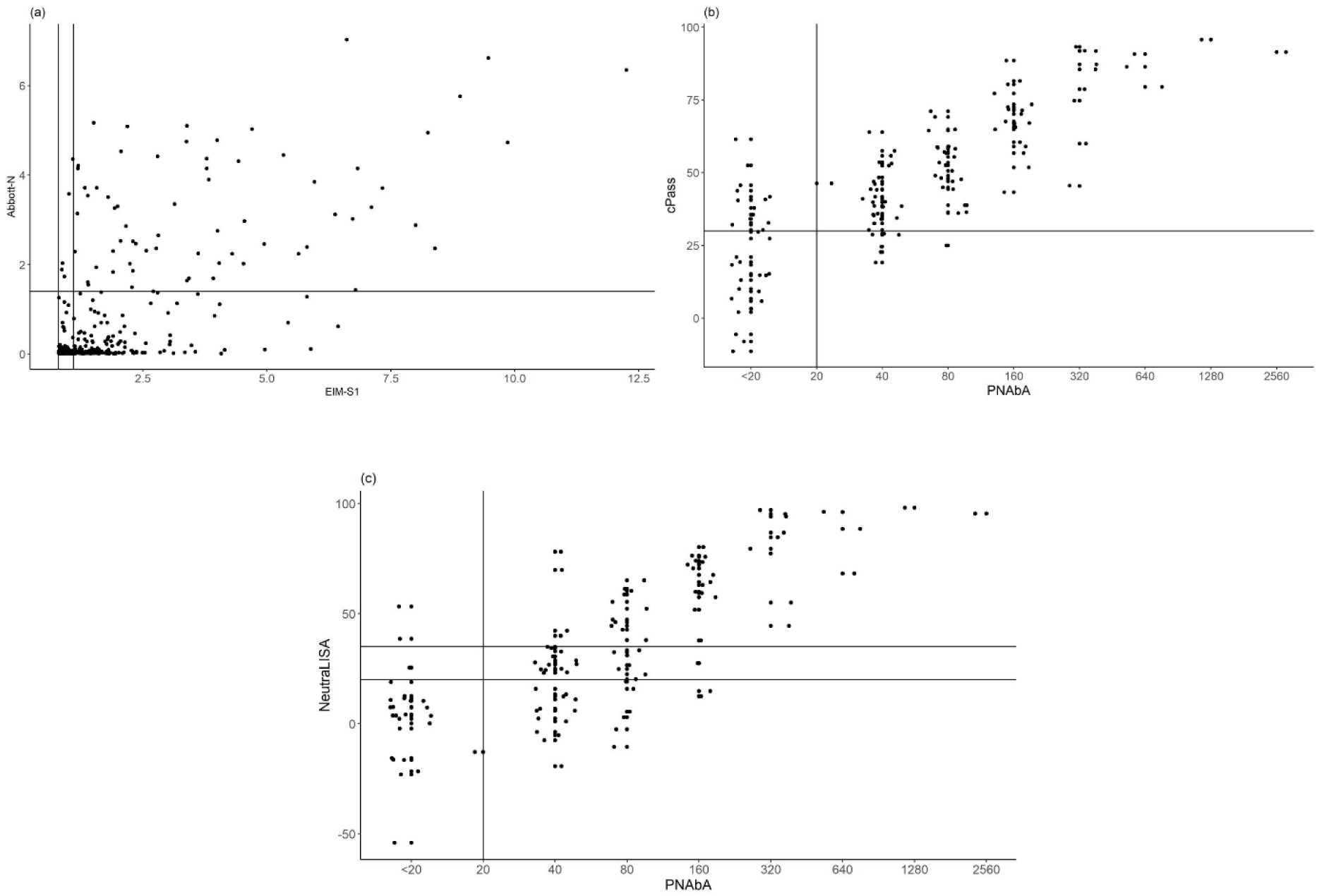
Comparison of EIM-S1 and Abbott-N (a) and PNAbA and cPass^TM^ (b) and NeutraLISA (c). PNAbA titres are expressed as log2-scale and 0.3 jitter has been used for clarity. Manufacturer’s cut offs are marked with vertical and horizontal lines.

Significant differences were detected between the two IgG assays. Only 28% of EIM-S1 positive samples were positive with Abbott-N (≥1.4), which is even lower than 44% (67/240) reported in the previous study [22]. 91% of IgG positive patients were also NAb positive and 67 % of NAb positive patients were also IgG positive.

## Discussion

We studied the development of the pandemic during 2020 in Finland using retrospectively collected anonymous samples from individuals of the same age and most likely without major underlying disease burden. This large Finnish serosurvey covers uniquely the whole year of 2020.

In early 2020, only highly targeted diagnostics were carried out as resources were targeted for COVID-19. Nevertheless, we had access to samples of anonymous pregnant women, providing enough samples per month (1000 per month) and we used these to retrospectively study the pandemic throughout the whole year 2020. Even though the data does not fully reflect the entire population in Finland, the dataset consisting of pregnant women was comprehensive enough for this kind of comparative study because age and gender factors are excluded and differences in behavioral patterns are likely smaller than in a dataset consisting of the entire population.

Most of the seroprevalence studies use a timeframe of a few weeks to a few months and methodology has varied greatly, complicating comparison between studies. A review on seroprevalence in Europe in 2020 described a lot of heterogenicity depending on the country, timeframe, and study group [23]. Reported seroprevalences varied between 0.7 and 45.3% in health care workers and between 0.24 and 23% in community studies. The majority of the studies detected no differences between genders whereas comparison between age groups gave conflicting results [23]. Just like community studies, studies on pregnant women have reported seroprevalences ranging from less than 0.5% (e.g., Estonia) to over 20% (e.g., Spain) during the first wave of the pandemic [24–30]. Eskil et al. noted a gradual rise from 0.5% to 5.7% in pregnant women until the end of 2020 [26] and Mattern et al. didn’t detect differences between women at delivery and general community, suggesting that seroprevalence in pregnant women may reflect that of the whole population [28].

The positive predictive value of the test (aka is the positive test result true mark of the disease) is affected by the sensitivity and specificity of the test, but also by the disease occurrence in the population in that moment. As the disease occurrence was very low in 2020, we stressed high specificity over high sensitivity. Therefore, a two-tier system, detecting both S1 and N antibodies, was selected to ensure we detect only true positives at the early stage of the pandemic when the disease was still rare. From January until end of March, we didn’t detect any COVID-19 IgG or NAb positive individuals among 3000 samples. From April until December, comprising 9000 samples from HUS, Eksote, and Kymsote regions, the numbers slowly increased from 0.7% to 1.3% (IgG) and from 0.9% to 1.4% (NAb). A community follow-up including both sexes and different age groups carried out by THL (Fig. S6) reported 0.5-5.5% seroprevalences in HUS region between April and December 2020 [3]. These numbers are a bit higher than in our study, but considering the different detection protocols and confidence intervals, prevalence among pregnant women in our study doesn’t seem to significantly differ from the prevalence detected in the community. It is also possible that during pregnancy people tend to be more careful and avoid exposures, leading to slightly lower seroprevalence. Compared to other studies on pregnant women and community studies, our seroprevalences are in the lower part of the spectrum [24–30], which is expected considering that diagnosed case numbers in Finland were also relatively low compared to several other countries (Fig. 2c-d).

The second aim in our study was to evaluate the commercial surrogate immunoassays (NeutraLISA, EIM and cPass^TM^, GenScript) for detection of NAbs. Several studies have indicated that surrogate neutralization assays show moderate to good correlation with titres of neutralization assays [6, 8, 31, 32]. Here, we used a positive PNAbA result as a real indication of detectable NAbs. Among the negative PNAbA samples (N=29), NeutraLISA gave 90% and cPass^TM^ 55% concordant results. Among the positive PNAbA samples (N=91), NeutraLISA gave only 49% and cPass^TM^ 92% concordant results. Notably, NeutraLISA gave a lot of borderline results which is not a desirable phenomenon in a diagnostic setting. Girl et al. tested a 100% specificity for NeutraLISA with pre-pandemic samples but, like us, lower specificity (82%) with whole dataset of negative individuals tested with microneutralization assay (consisting of partially vaccinated, pre-pandemic, and convalescent sera). They speculated that there may be some true positives or the NeutraLISA cross-reacts with non-neutralizing antibodies [7]. In addition, Girl et al. noticed a difference in NeutraLISA sensitivity between vaccinated sera and convalescent sera (86% for all samples combined and 69.3% for convalescent sera) as compared to microneutralization assay [7]. This might explain the lower number in our study, too, as only convalescent sera were used. Gradinger et al. tested sensitivities of cPass^TM^ and NeutraLISA to be 91% and 56%, respectively, as compared to neutralization test from convalescent patients [8]. Like us, Hoffman et al. noted a good specificity and lower sensitivity of NeutraLISA and good sensitivity of cPass^TM^, but also stated that manufacturer’s cut off values may sometimes need to be re-evaluated [32]. Tan et al. reported a sensitivity of 96.7% for cPass^TM^ and 83.3% for Abbott as compared to PCR results >21 days after symptoms [33]. El-Ghitany et al. who compared NeutraLISA to anti-SARS-CoV-2 QuantiVac IgG ELISA (EIM) determined that 77% of IgG positive samples were NAb positive but only 2.5% of NAb positive samples were IgG negative. They did not compare the tests to microneutralization assay [34]. In our data, 60% of IgG positive samples were positive in NeutraLISA. If we had considered borderline results positive like El-Ghitany et al., it would have been 81%. Olbrich et al. noticed a lower sensitivity with microneutralization assay (81%) than cPass^TM^ (96%) when compared to PCR-positive results in the past and suggested that cPass^TM^ might be more sensitive in detecting past infection than traditional microneutralization assay [35]. However, more studies would be needed to determine if samples that are positive with surrogate assays but negative with neutralization assays (commonly accepted golden standard) have true neutralizing capacity. Hence, surrogate tests with possibly higher sensitivity than neutralization assays, should be interpreted with caution in diagnostic settings when it is important to know if the patient has some immune protection or not.

Selection of the diagnostic tests depends on many factors, including the intended use. If surrogate NAb assays are used in diagnostics settings, false positives can be considered problematic. One of the major applications of these assays is monitoring the immunity status of patients, for example immunosuppressed patients, and evaluating the need for vaccine booster. With this in mind, more development and more optimizations may be needed for surrogate assays to reach the performance needed in diagnostic settings. More data are also needed to establish real life performance of these surrogate assays, including the performance after vaccination and hybrid immunity.

## Data Availability

All data produced in the present work are contained in the manuscript or its supplementary material.

## Acknowledgments

We wish to thank Leena Palmunen and Aurora Díaz Pérez for technical assistance and Eemeli Pettersson for help with preparing the maps.

## Financial statement

This study was funded by Helsinki University Hospital fundings (A.J., grant numbers TYH2021110, TYH2023102) and Jane and Aatos Erkko foundation (O.V.).

## Author contributions

Data curation: JV, AJ, EK; Formal Analysis: JV, AJ, SS, EK; Funding acquisition: TS, AJ, OV; Investigation: AJ, SS, EK; Methodology: AJ, SS, EK; Project administration: AJ, EK; Resources: TS, AJ, EK, OV; Software: JV; Supervision: AJ, EK; Validation: JV, AJ; Visualization: JV, AJ; Writing – original draft: JV, AJ; Writing – review & editing: all authors.

