## Supplementary for "SARS-CoV-2 infections among pregnant women, 2020, Finland – cross-testing of neutralization assays"

Supplementary material

Virtanen J[1,2,*], Korhonen E[1,2], Salonen S[3], Vapalahti O[1,2,3], Sironen T[1,2], Jääskeläinen AJ[3]

[1]Department of Veterinary Biosciences, Faculty of Veterinary Medicine, University of Helsinki, Helsinki, Finland

[2]Department of Virology, Faculty of Medicine, University of Helsinki, Helsinki, Finland

[3]HUS Diagnostic Center, HUSLAB, Clinical Microbiology, University of Helsinki and Helsinki University Hospital, Finland

Following tables are in a separate file

- Table S1: Basic information of all samples and EIM-S1 results
- Table S2: Geographical data from different municipalities and hospital districts included in the study
- Table S3: Samples chosen for further studies based on EIM-S1 result

**Table S4.** Basic statistics of age

| | Mean | Median | Range | SD |
|---|---|---|---|---|
| **Hospital district** | | | | |
| HUS | 31.74 | 32.0 | 15–61 | 5.13 |
| Eksote | 30.72 | 31.0 | 16–46 | 5.11 |
| Kymsote | 30.61 | 31.0 | 14–48 | 5.43 |
| **Month** | | | | |
| Jan | 31.90 | 32.0 | 18–53 | 5.14 |
| Feb | 32.08 | 32.0 | 17–54 | 5.22 |
| Mar | 32.08 | 32.0 | 15–48 | 5.30 |
| Apr | 31.92 | 32.0 | 17–61 | 5.29 |
| May | 31.73 | 32.0 | 17–46 | 5.20 |
| Jun | 31.50 | 31.5 | 16–47 | 5.22 |
| Jul | 31.56 | 31.5 | 17–45 | 5.04 |
| Aug | 31.48 | 31.0 | 16–51 | 5.10 |
| Sep | 31.54 | 31.0 | 18–47 | 5.13 |
| Oct | 31.34 | 31.0 | 14–58 | 5.08 |
| Nov | 31.19 | 31.0 | 17–48 | 4.97 |
| Dec | 31.28 | 31.0 | 15–46 | 5.16 |
| **IgG result*** | | | | |
| pos | 30.23 | 30.0 | 17–40 | 5.19 |
| neg | 31.64 | 32.0 | 14–61 | 5.15 |
| Total | 31.36 | 32.0 | 14–61 | 5.16 |
| **NAb result*** | | | | |
| pos | 30.29 | 30.0 | 17–40 | 5.17 |
| neg | 31.44 | 32.0 | 14–61 | 5.16 |

*Patients were considered positive for IgG if both EIM-S1 and Abbott-N were positive and NAb if there was a detectable titer (>20) in PNAbA.

**Table S5.** Spearman's correlation coefficiencies and Cohen's kappa values (and p-values) between each test

| **Spearman's rho (p-value)** | | | | |
|---|---|---|---|---|
| | EIM-S1 | Abbott-N | cPass | NeutraLISA |
| Abbott-N | 0.516 (<2.2E-16) | | | |
| cPass | 0.825 (<2.2E-16) | 0.448 (2.3E-7) | | |
| NeutraLISA | 0.847 (<2.2E-16) | 0.499 (1.5E-8) | 0.884 (<2.2E-16) | |
| PNAbA | 0.738 (<2.2E-16) | 0.519 (9.1E-10) | 0.819 (<2.2E-16) | 0.757 (<2.2E-16) |
| **Cohen's Kappa* (p-value)** | | | | |
| | EIM-S1 | Abbott-N | cPass | NeutraLISA |
| Abbott-N | 0.060 (0.048) | | | |
| cPass | 0.155 (<0.001) | 0.335 (<0.001) | | |
| NeutraLISA | 0.070 (0.001) | 0.377 (<0.001) | 0.265 (<0.001) | |
| PNAbA | 0.167 (<0.001) | 0.349 (<0.001) | 0.511 (<0.001) | 0.281 (<0.001) |

*0-0.2 is considered slight, 0.21-0.4 fair, 0.41-0.6 moderate, 0.61-0.8 substantial and 0.81-1 near perfect agreement

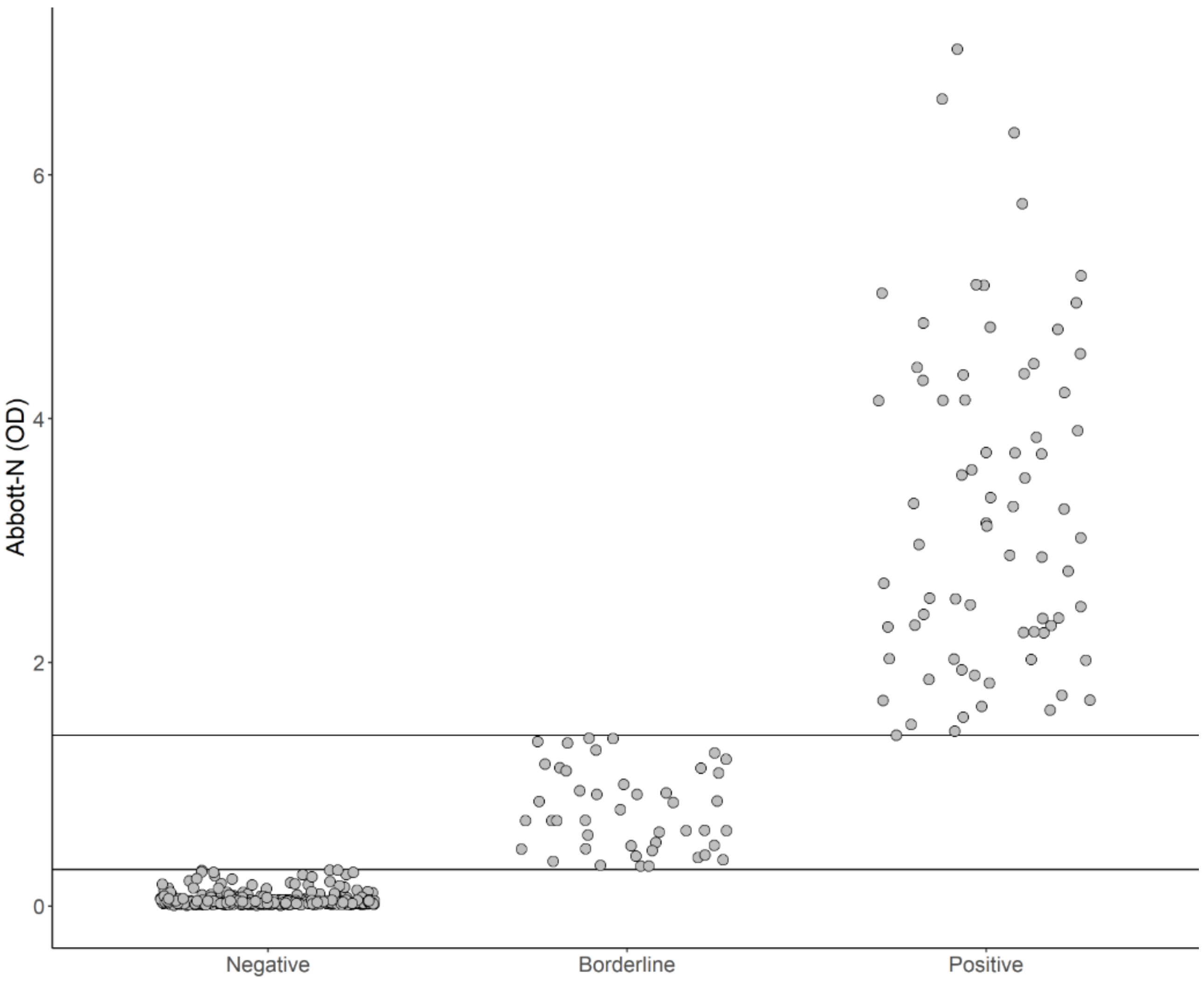


**Fig. S1.** Index values of samples tested with Abbott. Manufacturers limit of positivity (1.4 index) and limit for borderline result used in this study (0.3 index) are marked as horizontal lines. Jitter function (0.3) has been used for clarity.

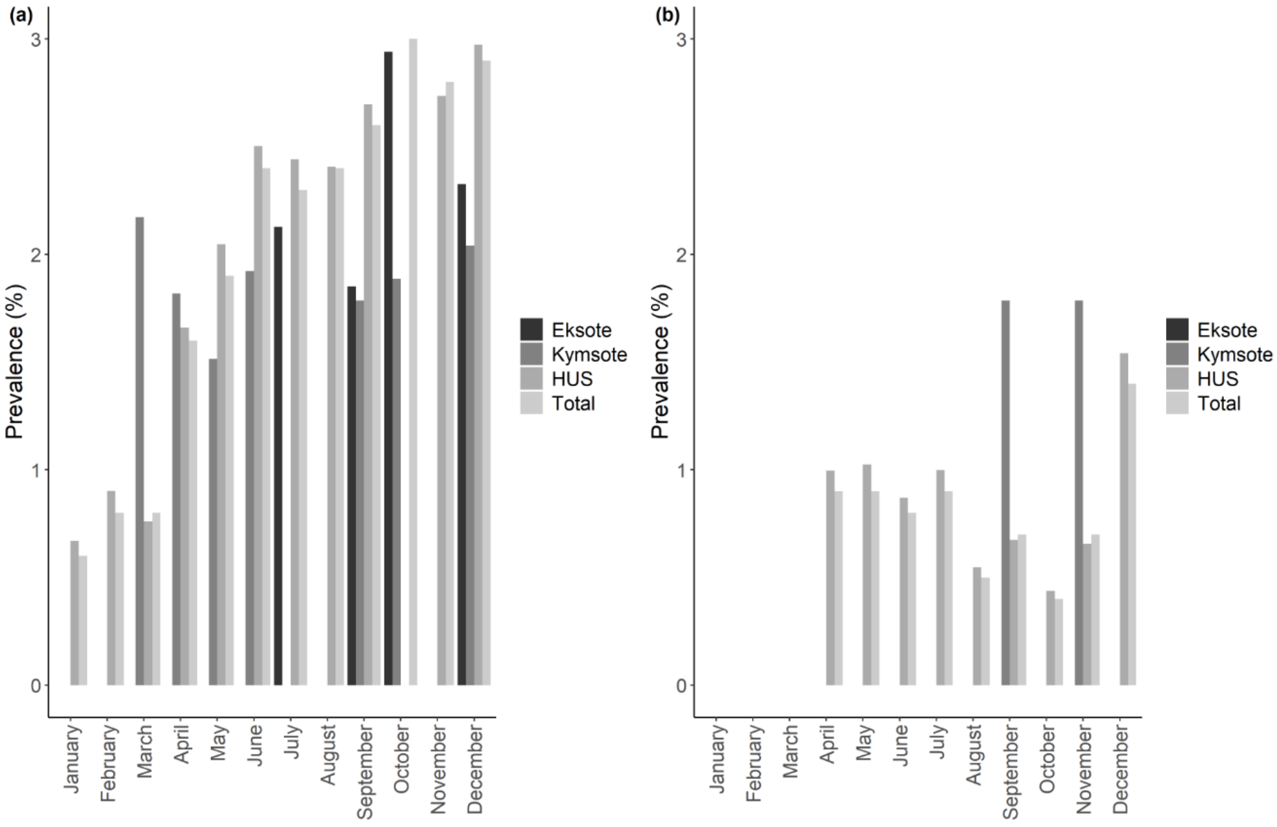


**Fig. S2.** SARS-CoV-2 IgG prevalence with EIM-S1 (a) and Abbott-N (b) in three hospital districts in Southern Finland in 2020. Cases that were positive with both IgG tests according to manufacturer's guidelines were counted as positives for IgG.

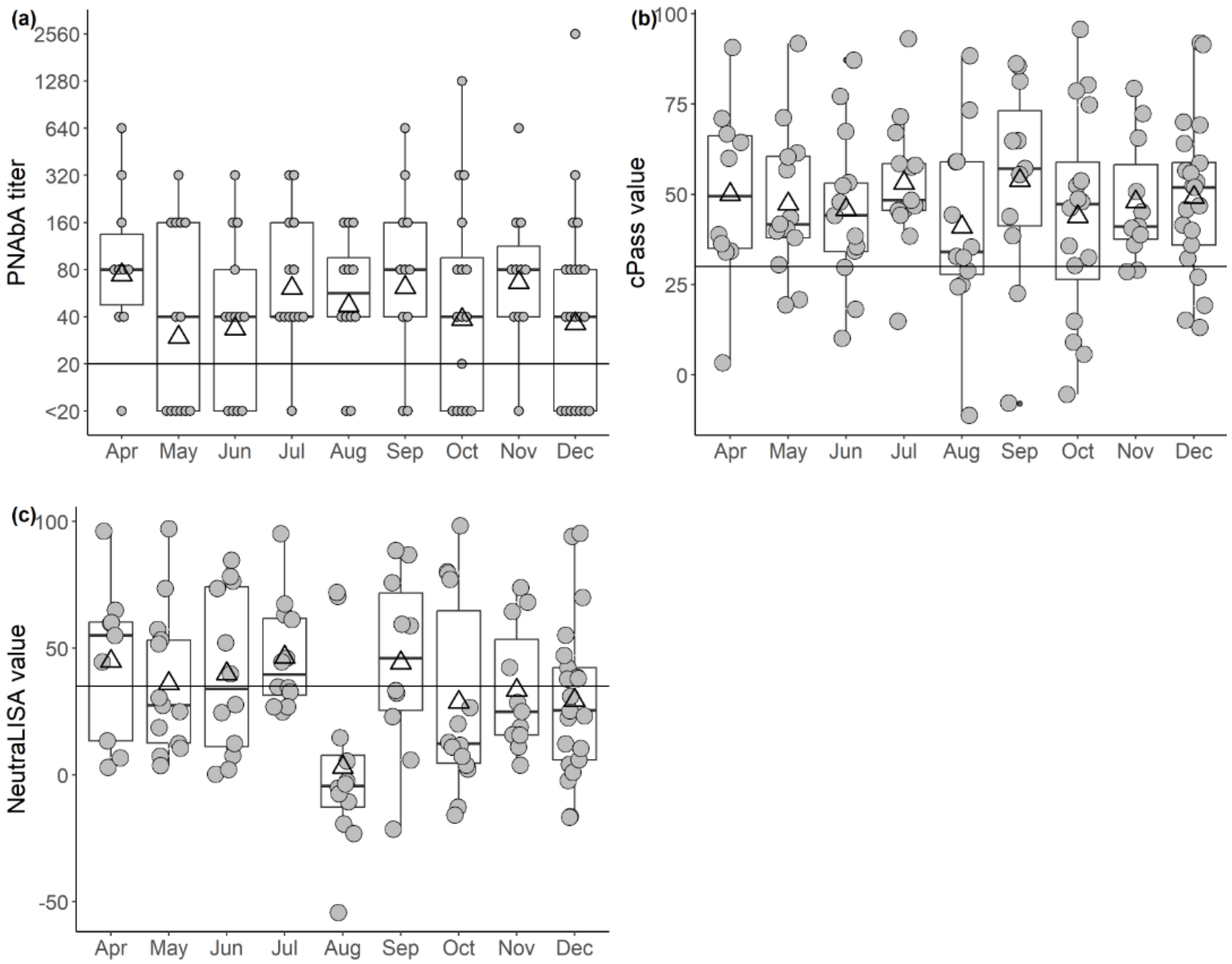


**Fig. S3.** Absolute values of PNAbA (a), cPass (b), and NeutraLISA (c). Limit of positivity is marked with horizontal line.

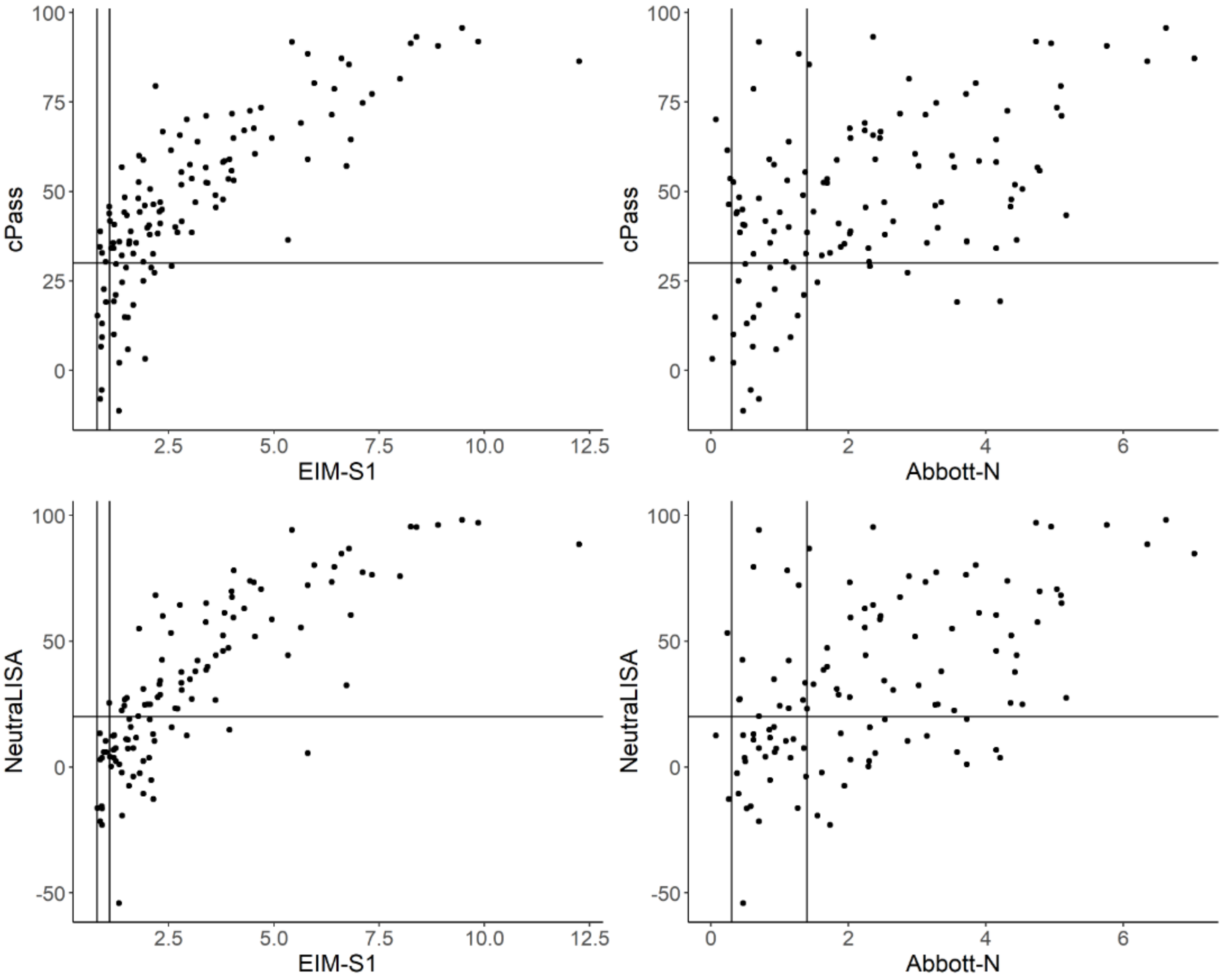


**Fig. S4.** Comparison of cPass and NeutraLISA to EIM-S1 and Abbott-N results. Manufacturer's cut off values are marked with horizontal and vertical lines.

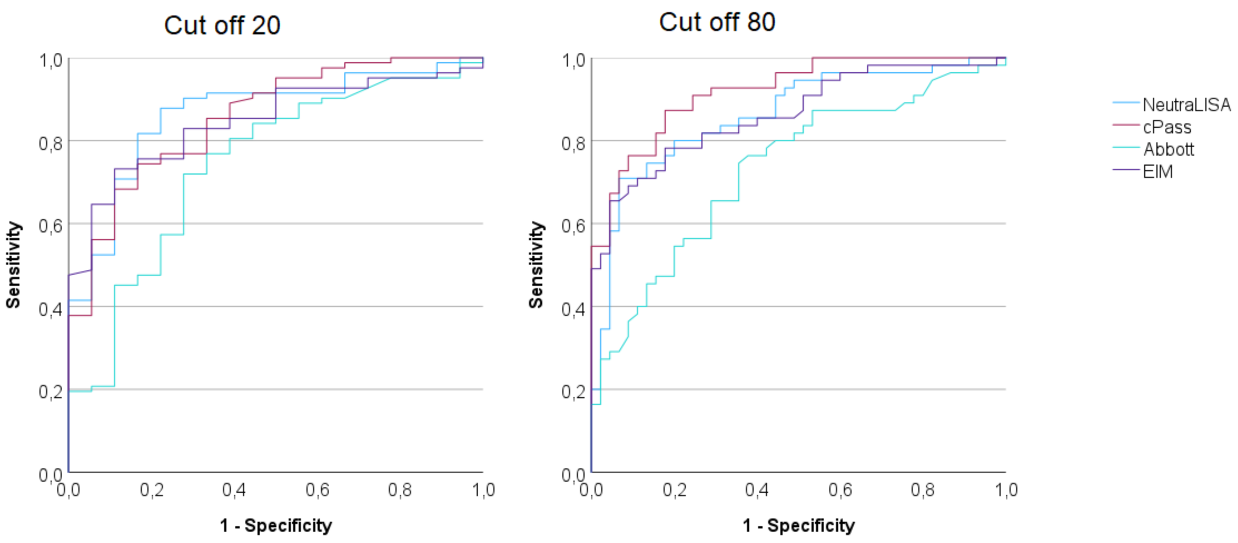


| | NAb titer cut off 20 | | | NAb titer cut off 80 | | |
|---|---|---|---|---|---|---|
| | AUC | Sensitivity | 1- Specificity | AUC | Sensitivity | 1- Specificity |
| cPass | 0.867 | 92 % | 50 % | 0.920 | 100 % | 71 % |
| NeutraLISA | 0.865 | 56 % | 11 % | 0.862 | 75 % | 16 % |
| Abbott-N | 0.758 | 70 % | 28 % | 0.732 | 81 % | 49 % |
| EIM-S1 | 0.882 | 96 % | 66 % | 0.864 | 98 % | 87 % |

**Fig. S5.** A ROC curve of all tests against PNAbA calculated using NAb titer thresholds of 20 and 80. The table shows individual AUC values as well as sensitivities and false positive rates (1-specificity) when manufacturer reported thresholds were used.

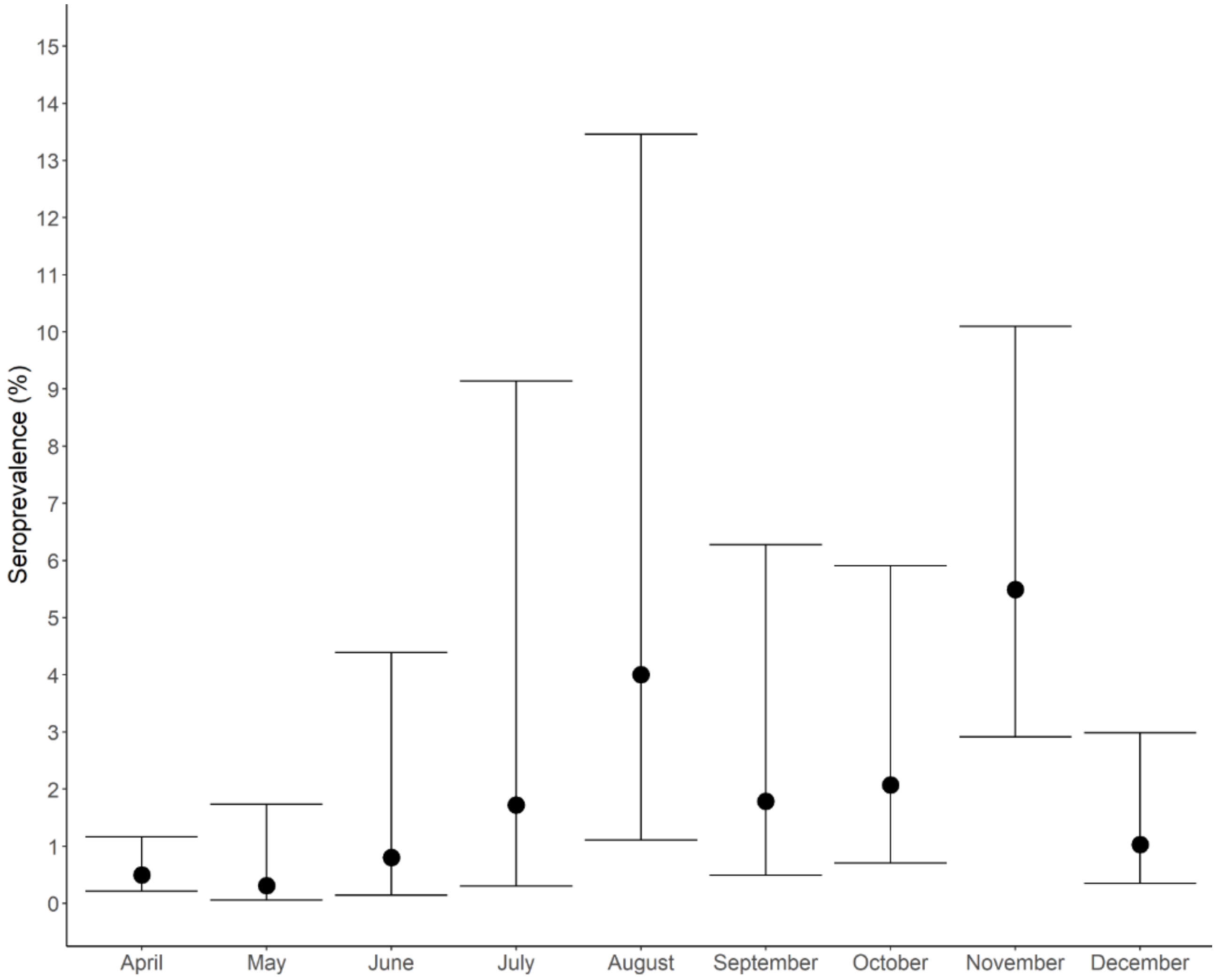


**Fig. S6.** SARS-CoV-2 community seroprevalences in Helsinki and Uusimaa hospital district in 2020 according to Finnish institute for health and welfare (Report of THL serological population study of the coronavirus epidemic. 2020. Available from: https://www.thl.fi/roko/cov-vaestoserologia/sero_report_weekly_en.html). 95% confidence intervals (Wilson Score Interval method) are included.
